# The magnitude of exercise-induced hypoalgesia can be improved and correlated with endogenous pain modulation following 4 weeks of treadmill running

**DOI:** 10.1101/2024.03.27.24304823

**Authors:** Zi-Han Xu, Nan An, Shuang Xu, Ru-Yun Wang, Yue Li

## Abstract

**Objective:** We aimed to investigate changes in pain perception, acute exercise-induced hypoalgesia (EIH), and endogenous pain modulation responses following 4-week treadmill running exercises of different intensities in healthy individuals.

**Methods:** Fifty-six healthy individuals included in this study. All participants were randomly assigned to three experimental groups (TRH, high intensity running, TRM, moderate intensity running and TRL, low intensity running). All participants performed 12 treadmill running sessions within 4 weeks at different intensities based on their target heart rate (THR). A running assessment was administered one week before running sessions. The magnitudes of EIH, conditioned pain modulation (CPM), and temporal summation (TS) responses following regular treadmill running were assessed. Pressure pain thresholds (PPT) or mechanical pain thresholds (MPT) were also determined following regular treadmill running.

**Results:** Treadmill running only induced acute EIH responses, with all pre-running PPT and MPT remaining unaltered. The acute EIH following each running sessions and CPM responses were also significantly improved in both the TRM and TRL groups, with TS score decreased in the TRM group.

**Conclusions:** A 4-week low to moderate intensity treadmill running improved acute EIH response by enhancing endogenous pain modulation in healthy individuals. Future studies should consider sex, behavior, and physiological factors to provide a comprehensive understanding of the changes in EIH following regular exercises.

## Introduction

Acute reduction of pain perception in the body following a single bout of exercise, commonly called exercise-induced hypoalgesia (EIH), has been widely confirmed in healthy individuals and some patients with pain[1]. Usually, both the global aerobic[2] and local resistance exercise[3] with a certain intensity and duration can temporarily increase various pain thresholds (pressure pain thresholds, PPT or mechanical pain thresholds, MPT) and enhance emotional well-being[4]. However, the EIH response may be weakened (absence of hypoalgesia or hyperalgesia) in older adults [5] or patients with painful conditions[6] and contribute to the impairment of endogenous pain modulation [7] (pain sensitization or pain-related psychological syndrome) in these individuals.

Exercise has been recommended as a non-pharmacological intervention and overall health promotion for various patients with pain and older adults, and the attenuation of chronic pain syndrome and improvement of pain-related behavior following regular exercise training, also known as training-induced hypoalgesia (TIH) have been reported in many studies [8-10]. However, there is no clear evidence proving that the magnitude of the analgesic effects following a single bout of exercise can be improved or restored by regular training in healthy individuals or patients with pain. [11]

In both healthy individuals and patients with pain, the EIH magnitude is affected by conditioned pain modulation (CPM) [12, 13] or temporal summation (TS) [14], which refers to the function of endogenous pain modulation and usually changes in individuals with sensitization of pain perception. In healthy individuals, pain perception can be inhibited or facilitated by descending control of the midbrain [15] and cortex [16] when the thalamus [17] receives certain inputs from peripheral nociceptors such as C fibers, which might also be activated by exercise with sufficient loads, leading to EIH. Therefore, it is important to understand whether endogenous pain modulation can be improved through regular exercise.

Additionally, the EIH magnitude can be modulated by the intensity of exercise [18]; high-intensity exercises often exacerbate pain in both human and rodent studies [19, 20], while moderate-intensity training increases the pain threshold in many conditions [21, 22]. Previous studies [23, 24] have shown that the relationship between EIH and exercise intensity is an inverted U-shaped curve in healthy individuals. Considering the possibility of pain exacerbation following high-intensity exercise [25], the persistent influence on pain perception and endogenous pain modulation may also differ between high- and moderate-intensity exercises.

Therefore, we aimed to compare the persistent effects of high, moderate and low intensity exercise on EIH and endogenous pain modulation in healthy individuals following 4-week high- and moderate-intensity exercises. We measured the effects of EIH in every exercise session and changes in CPM and TS responses before and after the 4-week training session. We hypothesized that (1) both high, moderate and low intensity exercises might elicit EIH responses (increase in PPT or MPT) in every exercise session, (2) the magnitude of EIH and CPM responses might gradually improve following regular low and moderate intensity exercise with the attenuation of TS response, and (3) the magnitude of EIH might be correlated with the CPM and TS responses.

## Methods

This study was approved by the Sports Science Experimental Ethics Committee of the Beijing Sport University (approval number: 2023023H) and was registered in the Chinese Clinical Trial Registry (registration number: ChiCTR2300074367).

### Study design

Altogether, 60 healthy participants included in this study performed exercise interventions 12 times within 4 weeks. All participants provided written informed consent. Demographic data and baseline measurements (resting heart rate [HRrest], PPT, MPT, and CPM responses) were collected. The maximum heart rate (HRmax) was estimated using the formula [26]: HRmax=202.5-0.53*age and the reserved heart rate (HRR) was calculated as HRR=HRmax-HRrest. Real-time HR was collected and recorded via the HR belt worn by the participants during running. The first running session was performed 48 h after baseline measurements to avoid potential long-lasting analgesic effects of the CPM test.

All participants were randomly assigned to three experimental groups (TRH, high-intensity treadmill running, TRM, moderate-intensity treadmill running and TRL, low intensity treadmill running) and single-blinded to treadmill running methods. Randomized sequences were generated using Excel software.

The TRH, TRM and TRL group performed treadmill running with 70%, 55% and 40% HRR, respectively. Running speed was determined in accordance with the target heart rate (THR) during baseline measurements. The primary outcomes of this study were PPT and PPT changes before and after exercise sessions (EIH responses). All participants performed a single exercise session once a day, three times per week for 4 weeks (Figure 1).

**Figure 1.**
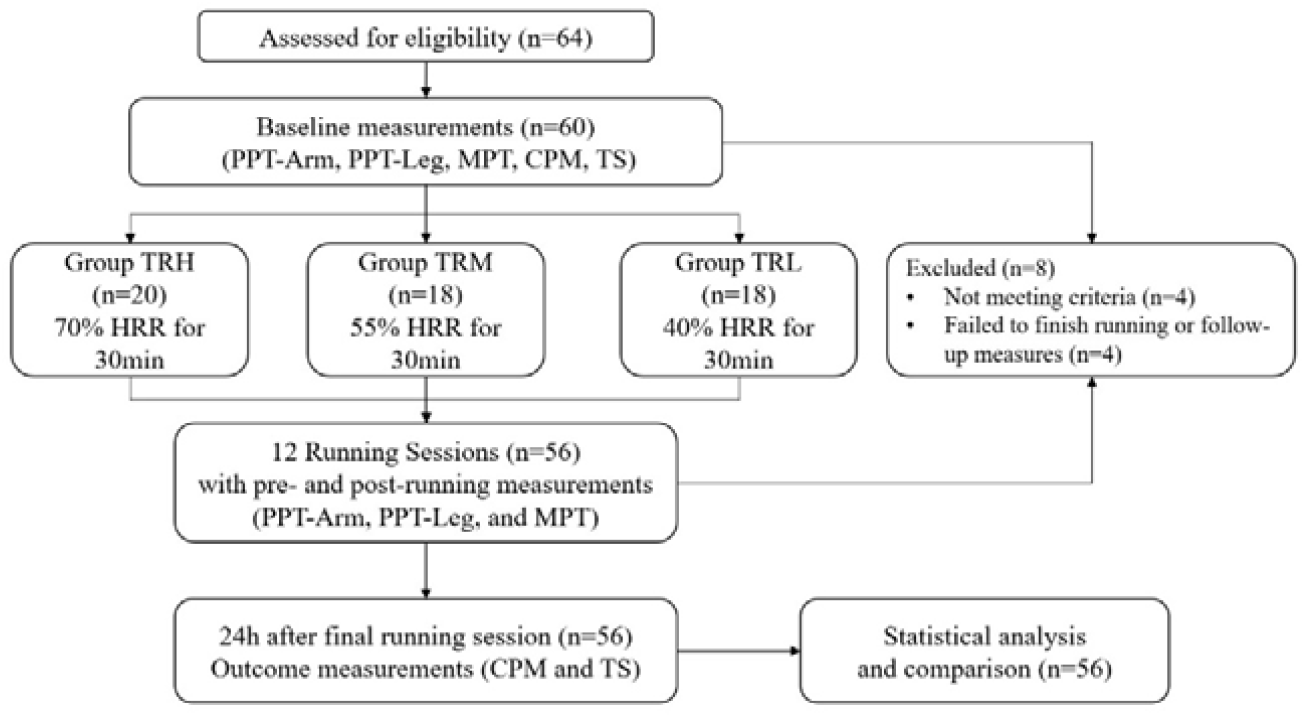
Flowchart of the experiment

### Participants

Based on previous studies [27], regular aerobic exercise-induced effect size on PPT changes ranging from 0.20 to 0.38 was 0.20. Our study utilized G*Power software with an effect size of 0.38, an alpha level of 0.05, and a power of 0.80. Thus, a minimum sample size of 48 participants across the three groups was determined.

Sixty-four healthy students (aged 18–30 years) from Beijing Sports University were assessed for eligibility in this study, of whom 60 were enrolled with 4 excluded. The exclusion criteria were (1) had pain-related pathological or psychological syndrome within 3 months; (2) had injury history of lower extremities within 1 year; (3) had potential or confirmed heart disease, or recovered from a heart disease <1 year ago, (4) failed to maintain or tolerate the exercise intensity during the regular treadmill running interventions; (5) showed serious exertion or fatigue in 24 h after any exercise session; (6) showed intolerable pain during the pain perception test; and (7) using combined hormonal contraceptives or any hormonal contraception (i.e., estradiol and progesterone) which may influence the pain perception. Besides, evidences[28, 29] suggests that the menstrual cycle does not have a significant effect on EIH, then participants who were menstruating would be included in this study.

### Procedures

All participants performed 12 treadmill running sessions within 4 weeks at different intensities based on their THR. The participants wore an HR belt to monitor and record real-time HR during the test and running sessions. A running assessment was administered to every participant one week before implementing the exercise interventions. This assessment involved a progressive increase in speed until the THR was reached. Subsequently, the predetermined speed for each individual was established at the commencement of running. During the running session, the running speed was adjusted at any time according to the participant’s heart rate changes.

### Outcome measures

Outcome measures were assessed at multiple time points, where the PPT-arm, PPT-leg, and MPT were recorded 5 min before and 10 min after each running session. As primary outcome, the evaluation of PPT was divided into two indicators: PPT-leg as local EIH responses; and PPT-Arm as global EIH responses. And the MPT were introduced to evaluate changes of mechanical perception as “sharp pain”. The CPM and TS responses were evaluated before and after all running sessions to determine the changes of individuals’ endogenous pain modulation function. All testing locations were marked with a sterile waterproof marker to ensure consistency in the repeated measures. The testing angle of the algometer was carefully adjusted perpendicular to the skin.

### PPT (Primary outcome)

The PPT was evaluated using a quantitative sensory testing protocol [30] with a handheld pressure algometer (Baseline Dolorimeter, Fabrication Enterprises, USA) equipped with a 1 cm^2^ metal probe. Pressure was applied at a rate of 0.5 kg/s over the right side of the two muscle groups: the extensor carpus radialis (PPT arm) and peroneus longus (PPT leg). The participants were instructed to indicate their perceived pain intensity using a visual analog scale (VAS) ranging from 0 to 100 cm. When the participants reported a pain intensity of 30 out of 100 cm (Pain30) during pressure application, the pressure thresholds were recorded as PPT values.

### MPT

The MPT was evaluated using a quantitative sensory testing protocol [31] with the handheld algometer equipped with a needle probe. Pressure was applied at 0.1 kg/s over the left side of the extensor carpus radialis. The participants were instructed to indicate their perceived pain intensity using the VAS ranging from 0 to 100 cm. When the participants reported a pain intensity of 30 out of 100 cm (Pain30) during pressure application, the pressure thresholds were recorded as MPT values.

### CPM

The CPM response was measured using a quantitative sensory testing protocol [30], specifically, the cold pressor procedure. Pressure was applied as the test stimulation, and cold-water immersion served as the conditioned stimulation. Participants first received pressure stimulation at the ipsilateral extensor carpus radialis, and the PPT was recorded as a test stimulus when the pain intensity reached Pain30. Subsequently, participants were instructed to immerse the contralateral hand into cold water at 8 □ for 1 min. The PPT at Pain30 was reassessed when the participants withdrew their hands from the immersion. The difference between the two PPTs was recorded in response to the CPM.

### TS

The TS response was measured using a quantitative sensory testing protocol [31]. The needle probe of the algometer was applied to the left side of the extensor carpus radialis at an intensity of 1.25 times the participants’ MPT. Subsequently, the mechanical stimulations were repeated 10 times at 0.5 Hz (1-second stimulus following a 1-second interval). Subsequently, the participants were instructed to report the pain perception of the first and last stimulations using the VAS score, and the differences in scores between these two mechanical stimulations were recorded as a response to the TS.

### Statistical analysis

The normality of all data was assessed using the Shapiro–Wilk test. Differences in baseline data (height, weight, HRrest, PPT, MPT, CPM, and TS) between both groups were analyzed using an independent t-test. The differences in the PPT and MPT values between pre- and post-running in each session were calculated as EIH responses, including EIH-A for changes in the PPT-arm, EIH-L for changes in the PPT-leg, and EIH-M for changes in the MPT.

A two-way repeated measures analysis of variance (MANOVA) was used to determine the differences between the three groups over running sessions and examine the EIH values of the PPT and MPT. And a one-way analysis of variance (ANCOVA) was also applied for between-group comparison of the CPM and TS responses, where the baseline results used as covariates.

The relationships between the CPM and TS, EIH-A, EIH-L, and EIH-M values after the running intervention were analyzed using the Pearson correlation method. All statistical analyses were performed using SPSS Version 21.0, and a significance level of p<0.05 was applied to all tests.

## Results

### Baseline characteristics

Four participants were excluded from this study because of myofascial pain syndrome that occurred one month before the experiments. Of the 56 participants enrolled in this study, 20 in the TRM group completed 12 moderate-intensity running sessions, and 18 in the TRH group completed 12 high-intensity running sessions. Additionally, two participants withdrew from the study because of the failure to finish all running sessions. No significant differences were observed in baseline characteristics between the groups (p > 0.05, Table 1).

**Table 1.** Baseline measurement (M±SD) ^1^.

|  | TRH (n=18) | TRM(n=20) | TRL (n=18) | P |
| --- | --- | --- | --- | --- |
| PPT-arm (kg/cm<sup>2</sup>) | 2.13±0.30 | 2.19±0.27 | 2.06±0.23 | 0.308 |
| PPT-leg (kg/cm<sup>2</sup>) | 3.98±0.48 | 3.83±0.42 | 3.96±0.32 | 0.475 |
| MPT (kg/cm<sup>2</sup>) | 0.62±0.16 | 0.57±0.10 | 0.57±0.07 | 0.270 |
| CPM (kg/cm<sup>2</sup>) | 0.58±0.23 | 0.64±0.30 | 0.61±0.22 | 0.814 |
| TS score (cm) | 28.42±6.10 | 31.99±6.40 | 27.50±7.67 | 0.103 |
| Age (y) | 21.22±1.93 | 21.30±2.77 | 21.17±2.04 | 0.984 |
| Height (cm) | 169.50±7.61 | 171.15±7.35 | 170.33±8.50 | 0.810 |
| Weight (kg) | 63.05±12.63 | 64.75±12.51 | 65.02±13.04 | 0.880 |
| Male/female | 6/12 | 8/12 | 7/11 | - |

All data were presented as mean ± standard deviation (M ± SD)
<sup>1</sup>: 1-way ANOVA, significant difference was set by $p \leq 0.05$ .

### Changes in EIH-A following running sessions

The two-way MANOVA revealed significant effects (p<0.001, F=9.424) for the running sessions involving the EIH-A, indicating that the 4-week running intervention significantly increased global EIH responses in all participants. The interaction effect (p<0.001, F=222.498) between running intensities and time for the EIH-A was also significant. However, post hoc tests showed that the EIH-A in the TRM group (p<0.001) since 2^nd^ running session, and in the TRL group (p<0.001) since 7^th^ running session were significantly higher than that in the TRH group, which indicated that only the global EIH in the TRM group improved after the 4-week treadmill running.

In addition, all pre-running PPT-arm values remained unaltered, indicating that regular running may not change the baseline level of the PPT-arm (Figure 2).

**Figure 2.**
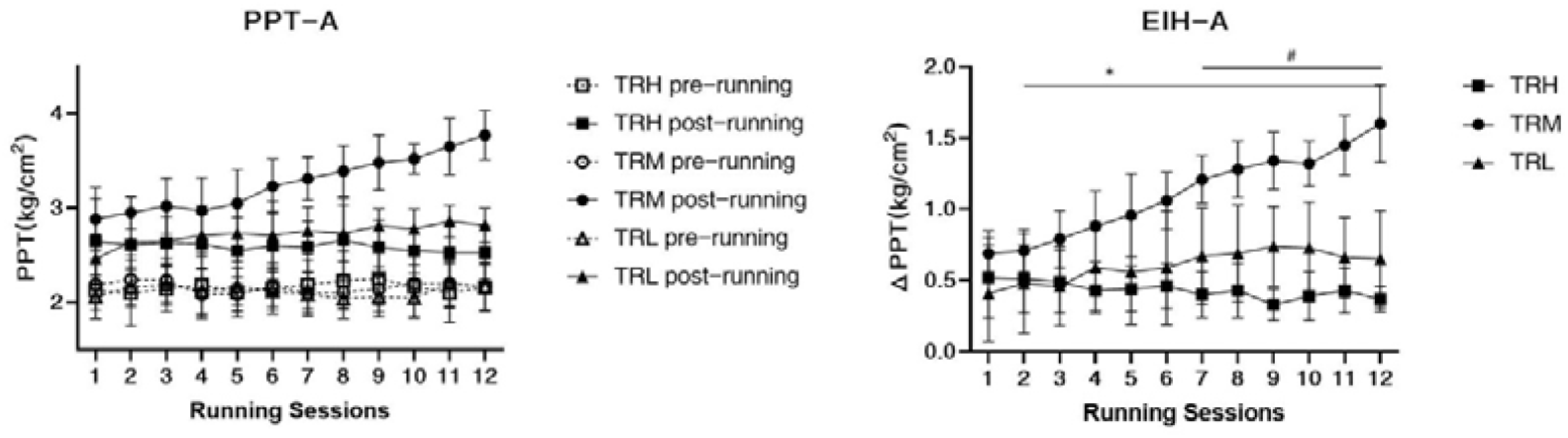
Changes in in PPT of arms and EIH-A following running sessions All data were presented as mean and standard deviation; PPT=pressure pain threshold; EIH=exercise-induced hypoalgesia; EIH-A=EIH value of PPT-arms; HRR=reserved heart rate *: EIH in TRM group significantly higher than TRH group ^#^: EIH in TRL group significantly higher than TRH group

### Changes in EIH-L following running sessions

Two-way MANOVA revealed significant main effects (p<0.001, F=6.909) for the running sessions involving the EIH-L, indicating that the 4-week running intervention significantly increased local EIH responses in all participants. The interaction effect (p<0.001, F=164.129) between running intensities and time for the EIH-L was also significant. However, post hoc tests showed that EIH-L in the TRM group (p<0.001) since 2^nd^ running session, and in the TRL group (p<0.001) since 7^th^ running session were significantly higher (p<0.001) than that in the TRH group, indicating that only the local EIH in the TRM group improved after the 4-week treadmill running. All pre-running PPT-leg values remained unaltered, indicating that regular running may not change the baseline level of the PPT-leg (Figure 3).

**Figure 3.**
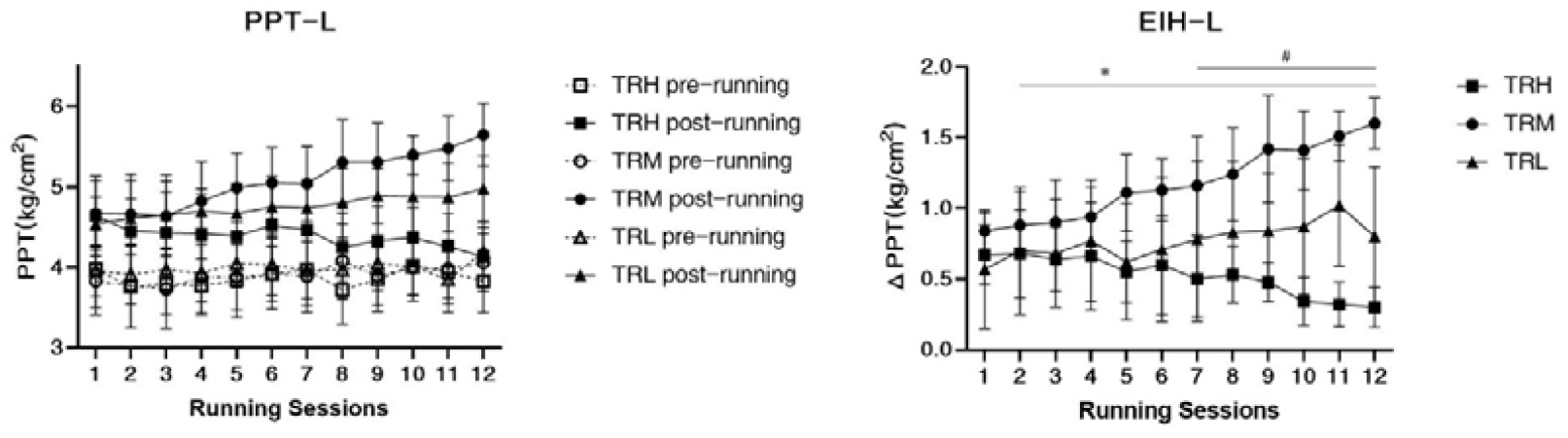
Changes in in PPT of legs and EIH-L following running sessions All data were presented as mean and standard deviation; PPT=pressure pain threshold; EIH=exercise-induced hypoalgesia; EIH-L= EIH value of PPT-legs; HRR=reserved heart rate *: EIH in TRM group significantly higher than TRH group ^#^: EIH in TRL group significantly higher than TRH group

### Changes in EIH-M following running sessions

The two-way ANOVA revealed significant main effects (p=0.004, F=2.084) for the running sessions involving the EIH-M, which indicated that the 4-week running intervention significantly decreased EIH responses in all participants. And there was no significant interaction effect (p=0.223, F=1.706) between running intensity and time for the EIH-M. Thus, ANCOVA test were applied and showed that the EIH-M in the TRH group was significantly higher (p<0.001) than that in the TRM and TRL groups in the 1^st^ to 3^rd^ running sessions. And EIH-M in the TRM and TRL groups were significantly higher (p<0.001) than that in the TRH group from the 9^th^ to 12^th^ sessions. This result indicated that the EIH-M in the TRH group gradually decreased following the 4-week treadmill running, whereas the EIH-M in the TRM and TRL groups was unaltered during the intervention. Additionally, all pre-running MPT values remained unaltered, indicating that regular running may not change the baseline level of the MPT (Figure 4).

**Figure 4.**
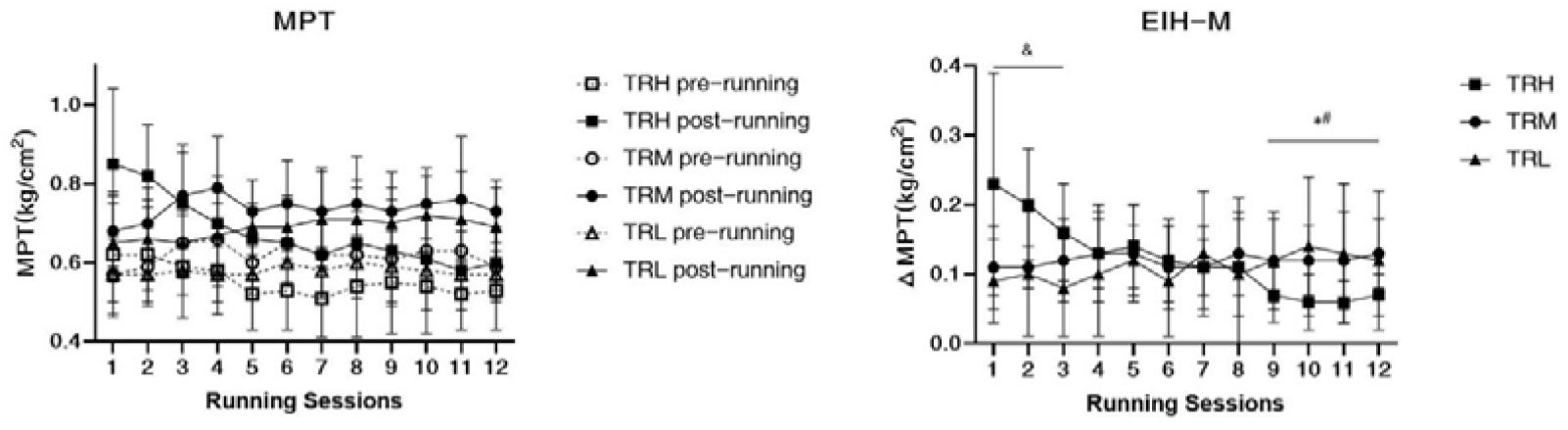
Changes in in MPT and EIH-M following running sessions All data were presented as mean and standard deviation; MPT=mechanical pain threshold; EIH=exercise-induced hypoalgesia; EIH-M=EIH value of MPT; HRR=reserved heart rate ^&^: MPT in TRH group significantly higher than TRM and TRL groups *: MPT in TRM and TRL groups significantly higher than TRH group

### Changes in CPM and TS following running sessions

One-way ANCOVA tests revealed significant between-group differences in the CPM (p<0.001) and TS responses (p<0.001) after the 4-week treadmill running intervention. The CPM responses in the TRM and TRL groups were significantly higher than those in the TRH group. In contrast, the TS scores of the TRM group were significantly lower than those of the TRH group, which showed no significant changes before and after the 4-week running intervention (Figure 5).

**Figure 5.**
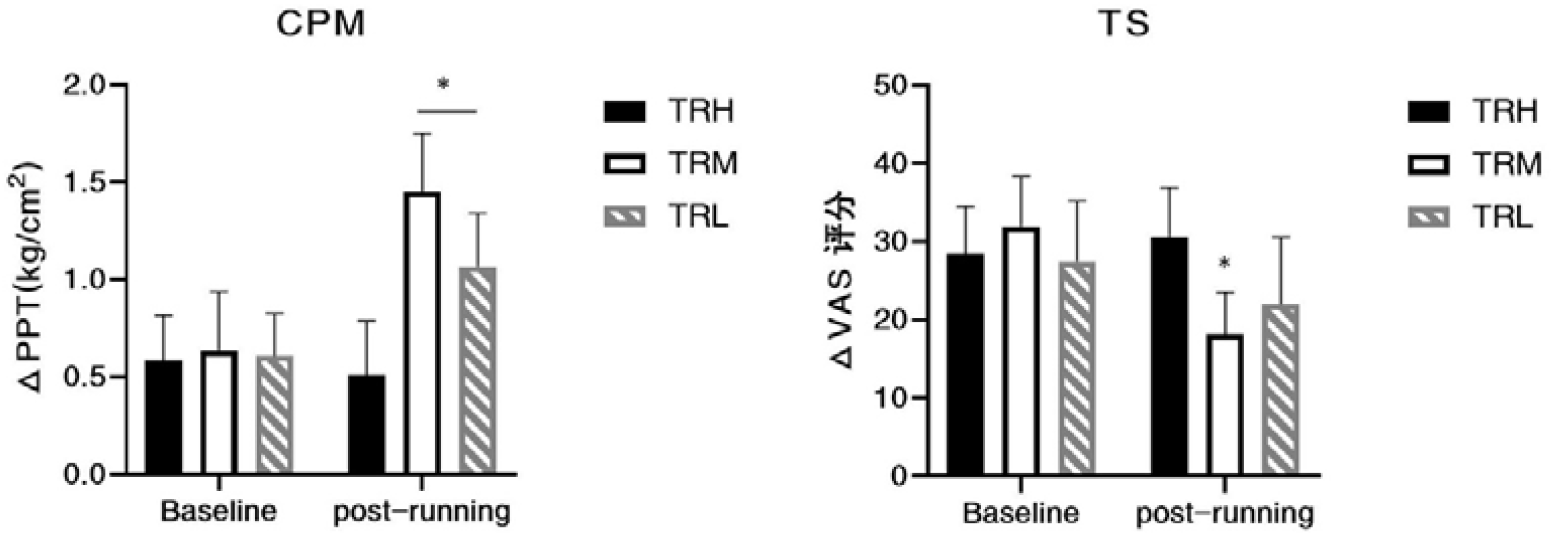
Changes in CPM and TS after 4-week running All data were presented as mean/standard deviation; CPM=conditioned pain modulation; *: significant difference compared with TRH group

### Relationship between endogenous pain tests and EIH magnitudes

Pearson correlation analysis includs all the results in the 12^th^ running sessions, and showed that there were significant positive relationships between CPM values and EIH-A (r=0.724, p<0.001), EIH-L (r=0.726, p<0.001), and EIH-M (r=0.347, p=0.009) magnitudes. There were also significant negative relationships between TS values and EIH-A (r =-0.529, p<0.001) and EIH-L (r =-0.544, p<0.001) magnitudes. However, the EIH-M (r =-0.209, p=0.121) magnitudes showed no significant relations to TS scores (Figure 6).

**Figure 6:**
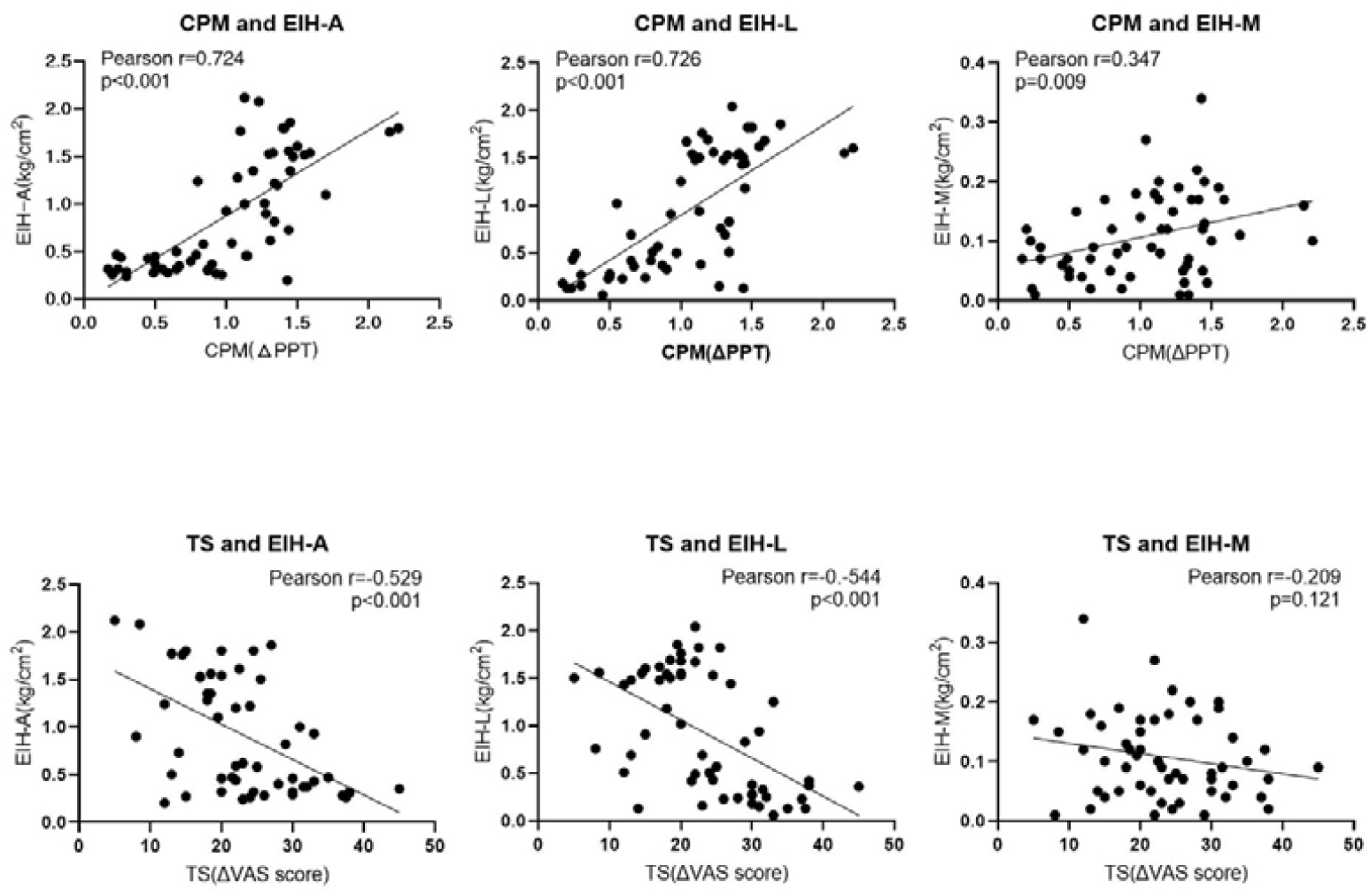
Relationship between CPM/TS and EIH magnitudes CPM=conditioned pain modulation; TS=temporal summation; EIH-A= EIH value of PPT-arms; EIH-L= EIH value of PPT-legs; EIH-M=EIH value of MPT.

## Discussion

We aimed to investigate changes in pain perception, EIH, and endogenous pain modulation responses following 4-week treadmill running exercises of different intensities in healthy individuals. Our results revealed the following: First, low- and high-intensity running may only induce short-term analgesic effects, including improvements in global and local PPT and MPT, within 24 h or less. Second, acute EIH responses following a running session varied according to the type of pain perception and exercise intensity, where the EIH-A and EIH-L following low and moderate intensity running were significantly increased along with exercise time, and the EIH-A and EIH-L following high intensity running slightly decreased after the exercise intervention. Third, the EIH-M following high intensity running significantly decreased with exercise time, and the EIH-M following low and moderate intensity running remained unaltered. Finally, the function of endogenous pain inhibition was enhanced, and facilitation was decreased following the 4-week moderate-intensity running exercise, showing positive and negative correlations with EIH responses, respectively.

The baseline pain perception threshold is relatively constant in healthy individuals [32] and may only be affected by activated or impaired endogenous pain modulation rather than regular exercise training. Recent studies have shown that a 24-week high-intensity interval training [33] and a 20-week resistant band exercise [34] have no significant effects on the PPT in healthy individuals. Tesarz et al. [35] investigated baseline pain perceptions in athletes and normally active individuals and observed that differences in pain thresholds between the groups were not significant.

However, as a response to endogenous pain modulation, EIH following exercise may change after a regular exercise intervention. Song et al.[11] suggested that exercise training induces physiological changes leading to improved EIH. Ohlman et al.[5] observed a greater EIH response in individuals who performed moderate physical activity per week than in sedentary controls. Hansen et al.[36] also observed that the PPT and EIH in healthy individuals significantly increased after a 7-week military training. However, evidence from randomized trials with a pre-test-post-test design remains limited.

Exercise with a sufficient load can induce a short-term EIH response, whereas exercise with low or moderate intensity may elicit greater analgesic effects than high-intensity or exhaustive exercise [23, 24]. Running at low- or moderate-intensity may activate non-noxious C fibers [37] via repeated muscle contractions, induce descending inhibition, upregulate 5-HT receptors in the brainstem [38], and attenuate pressure or thermal pain perception. High-intensity exercise may trigger noxious [39] and non-noxious C fibers and potentially induce descending facilitation [40] with limited EIH responses. Additionally, the upregulation of cannabinoids and opioids expression following high-intensity exercise may decrease the perception of mechanical stimuli. [41]

Therefore, we hypothesized that regular exercise may induce plastic changes in endogenous pain modulation. Low or moderate intensity exercise may enhance the central descending inhibition function and increase acute EIH responses with an increase in CPM responses. Lemley et al.[12] investigated EIH in healthy individuals and observed that those with greater CPM were more likely to experience greater EIH. Naugle et al.[42] also observed that healthy adults who self-reported increased total physical activity exhibited reduced TS and greater CPM.

In contrast, high intensity exercise may induce the adaptation of endocannabinoids and opioid modulation, decreasing the EIH of the MPT with unaltered baseline pain perception. For example, athletes experiencing high-intensity training showed a partially decreased EIH response than did healthy controls. Siebers et al.[41] observed a downregulation of endocannabinoid levels following regular running training. However, differential changes in the PPT, MPT, and other pain perception measurements represent various pain modulation pathways.

Our study had several limitations. First, the indicators of the pain tests were limited. For instance, adding heat pain detection thresholds might provide a more complete description of the changes in pain perception. Second, the intervention period for running exercise was relatively short. Future studies should investigate the long-term (> 8-12 weeks) effects on pain perception and modulation following various types of exercises. Finally, the male/female ratios in groups were not balanced. Considering the potential sex differences in endogenous pain modulation function and exercise behaviors, future studies should consider sex, behavior, and physiological factors to provide a comprehensive understanding of the changes in acute EIH following regular exercise.

## Conclusion

In summary, a 4-week low to moderate intensity treadmill running improved acute EIH responses by enhancing endogenous pain modulation in healthy individuals. CPM and TS may be correlated with EIH and changed after regular exercise, indicating that treadmill running may induce TIH through functional changes in endogenous pain modulation. However, baseline pain thresholds may remain unaltered and may not be affected by long-term exercise interventions.

## Acknowledgments

We would like to thank Tian-Rui Wu, Zheng-Quan Shi, Meng-Fei Lei, Zhao-Xia Zhou, Hua-Lian Tang, and all the researchers who provided assistance and advice during our experiments. And we would also like to thank Editage (www.editage.cn) for English language editing.

## Author Contribution

Zi-Han Xu, Nan An, Shuang Xu and Yue Li conceived and designed research, Zi-Han Xu, Nan An, and Ru-Yun Wang performed experiments, Zi-Han Xu and Shuang Xu analyzed data. Zi-Han Xu, Nan An, Shuang Xu and Ru-Yun Wang interpreted results of experiments. Zi-Han Xu and Yue Li prepared Tables and Figures and drafted manuscript. All authors edited and revised manuscript drafts and approved final manuscript.

## Data availability statement

Data available on request from the authors

